# Multiplex Portuguese Families as a Lens into rare mutations and the Shared Genetic Architecture of Schizophrenia, Mood Disorders, and Autism Spectrum Disorders

**DOI:** 10.64898/2026.04.06.26350177

**Authors:** Carlos N Pato, Michele T. Pato, Jennifer Mulle, Ronald P. Hart, Zhiping Pang, James A. Knowles, Tarjinder Singh, Priya Maddhesiya, Celia Carvalho, Alison Merikangas, Helena Medeiros, Tim B. Bigdeli, Hamed Kazemi, John Drake, Vladimir I. Vladimirov, Brion S. Maher, Silviu-Alin Bacanu, Ben Neale, Ayman Fanous

## Abstract

In an analysis of **173 multiplex families** from the **Portuguese Island Collection (PIC)** this study characterizes the shared genetic architecture of **serious mental illnesses (SMI)** including **schizophrenia (SZ)**, **bipolar disorder (BP)**, **major depression (MDD)**, and **autism (ASD)**. Within this cohort, co-segregation of psychotic and mood disorders occurred in 28% of families, while 7% demonstrated co-segregation of intellectual disability or ASD with SZ and mood disorder phenotypes. Whole-genome sequencing (WGS) was performed on a **three-generation PIC family** to identify rare, large-effect variants. This led to the identification of an extremely rare predicted **loss of function (LoF) mutation** in the ***Chromodomain Helicase DNA Binding Protein 2* (CHD2)** gene. These results demonstrate that high-density multiplex families in founder populations are a powerful resource for mapping rare, large-effect variants that cross clinical diagnostic boundaries, as the identified *CHD2* mutation suggests that the disruption of a single neurodevelopmental gene may lead to diverse SMI phenotypes. By combining population and family-based methodologies, this approach leverages shared genetic backgrounds and environments to provide a unique opportunity for cellular studies to explore the biological mechanisms underlying SMI, offering significant potential to inform future functional research and identify novel therapeutic targets.

## Introduction

Serious mental Illnesses (SMI) are devastating and chronic disorders that affect a significant portion of the population at tremendous individual, family, and societal cost. Recent findings point to overlapping and shared genetic architecture implicating both common and rare variants. There is now abundant evidence from large scale GWAS studies for **common** polygenic architecture underlying multiple psychiatric disorders, with schizophrenia and bipolar disorder in particular sharing common pleiotropic loci (1–4).

Studies of rare variants for both schizophrenia and bipolar disorder also implicate shared genetic factors underlying both conditions (5, 6). Collaborative studies by our team and others now implicate >50 rare genetic variants with high penetrance for SZ (7). Our parallel collaborative study of bipolar disorder revealed AKAP11 as a definitive shared risk gene with schizophrenia (OR ≈ 7.06, P ≈ 2.8 × 10⁻⁹) (6). Ongoing larger follow-up studies are increasing the number of identified rare variants, including rare Copy Number Variations (CNVs), that continue to demonstrate significant shared genetics for forms of schizophrenia, bipolar disorder, and autism spectrum disorders. These rare variants have large effect sizes (odds ratios), with each individual variant increasing risk by at least 3- and up to 100-fold. This high risk for phenotype at the macro-level of behavior suggests a similarly large effect at the micro-level of molecular and cellular perturbation of these SMI-associated genetic risk factors. A growing number of rare high risk genetic variants for SMI often present with shared symptoms and diagnoses. These similarities suggest the compelling hypothesis that *the mechanism of action of these variants may converge onto a limited set of pathways, cellular systems, or circuits*.

Studies within family pedigrees minimize genetic heterogeneity and population stratification that remain important challenges in genetic studies. Moreover, family-based studies often have enhanced diagnostic validity based on more extensive phenotyping than is generally pursued in large case-control GWAS studies. A meta-analysis of schizophrenia risk among relatives in a multiplex family reveals that in a family with one affected member risk for first degree relatives is eight-fold higher and increases to 11-fold in families with two affected members (8). This suggests that the genetic basis in some multiplex families is likely to be both inherited and represent more penetrant forms of illness.

Our study, the Portuguese Island Collection (PIC), was designed “…to limit the potential genetic heterogeneity of our study sample and select a population that was geographically isolated and was historically relatively genetically homogeneous” (9). Participants were ascertained as a population-based sample largely in the relatively isolated Azores and Madeira islands. These once-unpopulated islands were settled 600 years ago almost exclusively by the Portuguese. This PIC has been an integral part of several international collaboratives (e.g., the Psychiatric Genomics Consortium (PGC)).

Whereas many studies have focused on a specific diagnosis (i.e., schizophrenia, bipolar disorder, autism), the PIC was designed to use a whole population approach and ascertain all multiplex families defined as any family with at least two members with either psychosis or bipolar disorder. Importantly, the PIC includes systematic follow-up of the families since its inception in the 1990s, and we have identified new onset of illness in family members in the fourth generation. We have identified and enrolled all consenting probands and relatives on the Azores and Madeira.

At the time these studies were established, ASD was considered an exclusion for that individual in terms of a study focused on SZ and BP. Given these inclusion criteria, we included family and medical history for these family members, but they were not included in our genetic study. This may have led to an underestimate of ASD especially for mild cases and diagnostic definitions have become much broader over time. We also have minimal data for ASD in the general population in general only identifying severe cases in these multiplex families. However, given the evidence for genetic overlap between ASD and SZ, we have examined our PIC families for co-segregation with ASD.

## Results

In the PIC cohort, a total of 173 families with 2 or more affecteds were identified with either SZ or BP. Psychosis was present in most families. In 61 families (35% of the PIC families) all affected members were diagnosed with SZ. In 63 families (36% of the PIC families) all affected members were diagnosed with bipolar disorder or other mood disorders, with or without psychosis. Though psychosis was present in most families, most of the families segregated either mood disorder or schizophrenia spectrum diagnoses. However, 49 families (28%) presented with multiple relatives carrying a range of diagnoses including schizophrenia, schizoaffective disorder, bipolar disorder and major depression (with or without psychosis). Many of the more densely affected pedigrees segregated a variety of diagnoses. (Figure 1).

**Figure 1:**
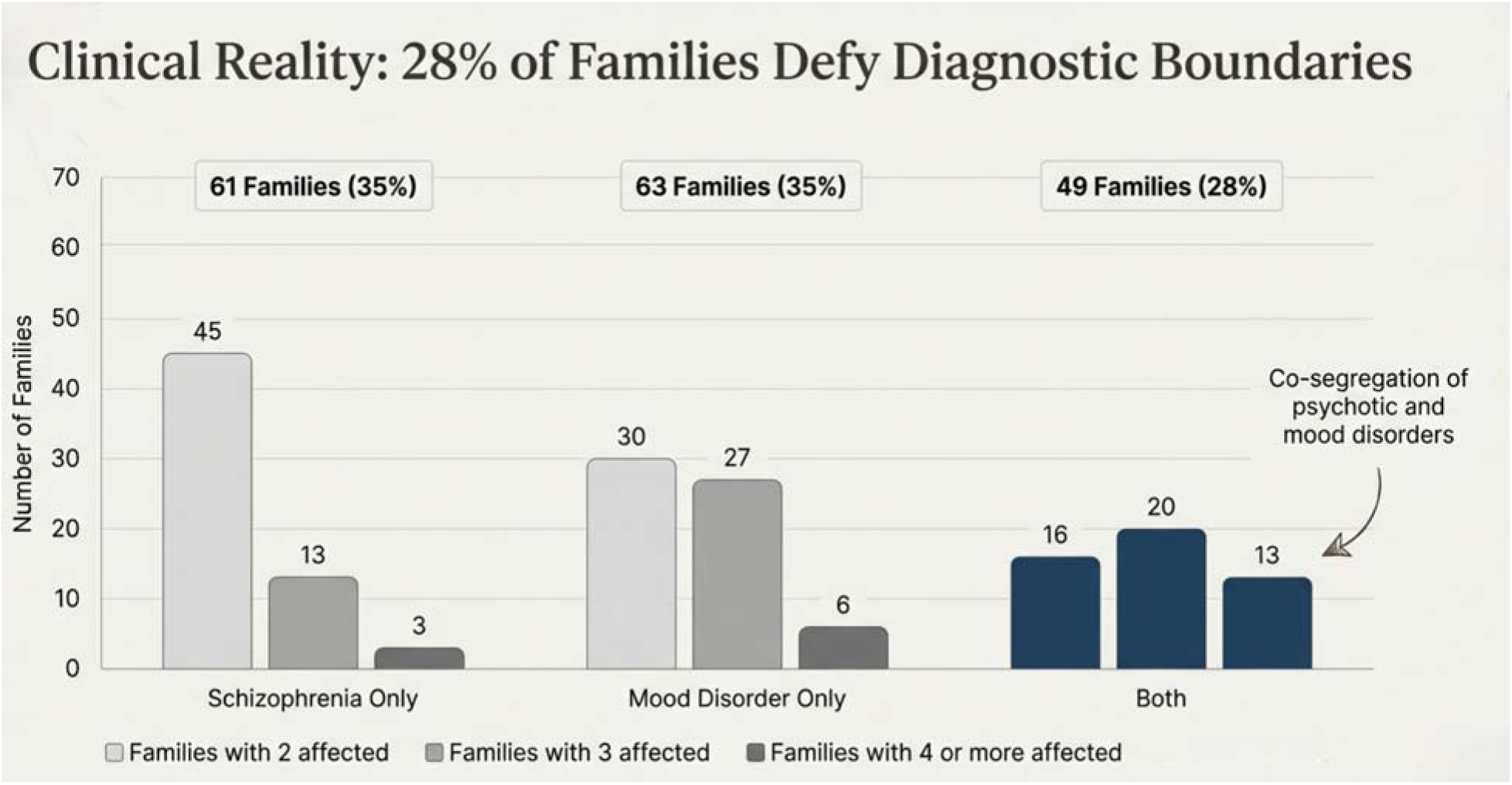

The number of affected family members in each of the PIC multiplex families range from 2 to 11. For families where all affected members carry the diagnosis of SZ 26% have 3 or more affected members. For families where all affected members carry a mood disorder diagnosis 52% have 3 or more affected members. Whereas, for families co-segregating both SZ and mood disorders 67% have 3 or more affected members.

The PIC families also include 12 families that co-segregate ASD/ID with SZ and or mood disorders. These 12 families were ascertained through an initial proband carrying the diagnosis of either SZ or BP. The characteristics of these twelve families are presented in table 1. Eleven of the twelve families have both SZ and ASD/ID affected family members, of these three also have members suffering with mood disorders, and one co-segregates bipolar disorder with psychosis and ASD/ID.

**Table 1:** PIC Families co-segregating ASD/ID.

| Family ID | Primary Diagnosis | NDD Phenotype | Pedigree Density |
| --- | --- | --- | --- |
|  |  |  | (Affected n) |
| Family A | SZ | ASD + ID | 5 |
| Family B | SZ | ASD + ID | 4 |
| Family C | SZ | ASD + ID | 3 |
| Family D | SZ | ASD + ID | 6 |
| Family E | SZ | ASD + ID | 5 |
| Family F | SZ | ASD + ID | 3 |
| Family G | SZ | ASD + ID | 2 |
| Family H | SZ | ASD + ID | 2 |
| Family I | SZ/BP | ASD + ID | 6 |
| Family J | SZ/BP | ASD + ID | 6 |
| Family K | SZ/BP | ASD + ID | 4 |
| Family L | BP with Psychosis | ASD + ID | 9 |

As a trial of screening for rare variants in multiplex families, we identified **Family A** (Figure 2) that co-segregates schizophrenia and ASD. Four family members across three generations were whole genome sequenced (WGS).

**Figure 2:**
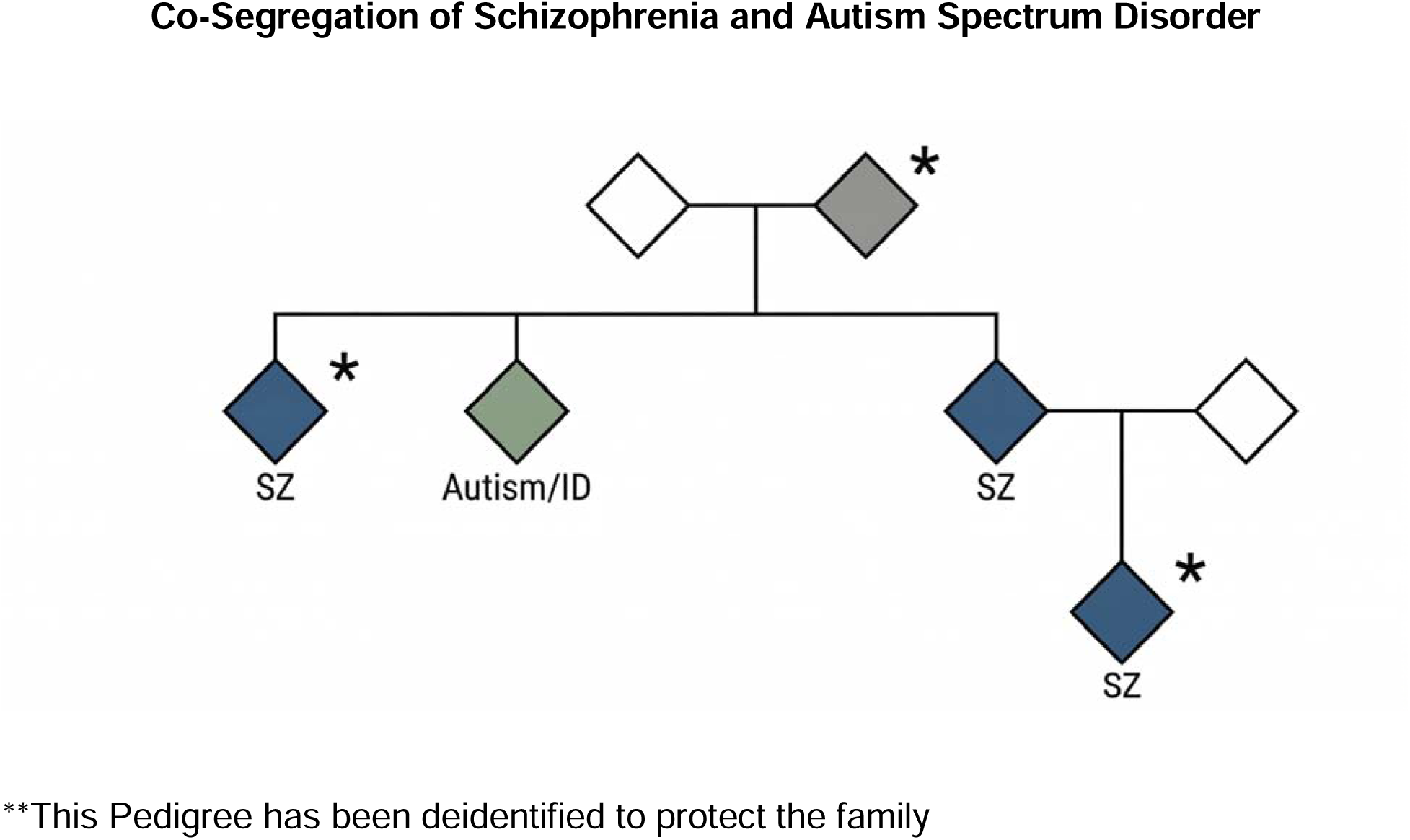
Family A.

We screened for rare predicted LoF and known psychiatric mutations in neurodevelopment genes and discovered an ultra-rare stop-gain predicted LoF mutation in Chromodomain Helicase DNA Binding Protein 2 **(*CHD2*)**, a SFARI Category 1 gene for autism (10). The variant is at chr15:93024653 C-A and did not appear in the Schizophrenia exome meta-analysis consortium (SCHEMA) database nor the Bipolar Exomes (BiPEX) variants list in either cases or controls. It appeared once in over 800,000 unrelated individuals in gnomAD v4.1(11). The family has multiple affected members; a grandparent was diagnostically assessed and may have a milder non-psychotic phenotype. Several surviving progeny are affected, and one third generation family member is affected. One 2^nd^ generation family member carries the diagnosis of Autism (ASD)/ Intellectual disability (ID) by history. All the other affected family members carry the diagnosis of schizophrenia and were formally diagnosed by our team. One family member has been formally diagnosed with schizophrenia but does not carry the CHD2 variant.

## Discussion

Our case control based Genomic Psychiatry Cohort (GPC) study with over 65,000 participants, has identified a number of cases carrying rare highly penetrant variants (12). Several hundred affected participants carry known CNVs and predicted LoF mutations that are significant in SCHEMA and BIPEX. These patients were identified using narrowly defined inclusion criteria.

Studies of CNVs show that each manifest a variety of clinical phenotypes ranging from schizophrenia and bipolar disorder to autism and developmental delay; furthermore, many individuals may have more than one diagnosis. For example, individuals with the 3q29 deletion have a roughly 40-fold increase in the risk of developing schizophrenia compared to the general population, and they may also have mild cognitive disability, ASD, anxiety disorders, and ADHD (13, 14). Individuals with 22q11.2 deletion syndrome may also exhibit a wide range of neurodevelopmental and psychiatric morbidity, including high risk for schizophrenia. However, there are multiple reports of people with 22q11.2 deletion syndrome escaping genetic diagnosis until well into their adult years (15, 16). This serves as a cautionary note since carrying one of these clearly high-risk rare variants may not lead to a clinically definable SMI phenotype.

Analyses of multiplex families from the PIC study showed co-segregation of psychosis and mood disorders, as well as other diagnoses. We hypothesize that one mechanism that might account for multiplex families is the presence of highly penetrant rare variants that may be identical by descent and follow a complex pattern of inheritance. These rare variants carry a high odds ratio for developing illness and likely represent a rare form of illness. For example, Family A appears to be the first multiplex family reported to segregate a *CHD2* predicted LoF mutation to present with schizophrenia as the most common phenotype. One sib carries the diagnoses of intellectual disability and severe autism. This may represent a family where the rare variant can present as different clinical syndromes.

The presence of a family member, who we formally diagnosed with SZ and lacks the ***CHD2*** mutation, identifies them as a phenocopy and suggests a high-density multiplex architecture beyond a single large-effect rare variant. Multiplex families are likely to carry significant polygenic risk and/or environmental risk burden. Ultimately, this pedigree supports a “multi-hit” model where the ***CHD2*** mutation acts as a potent modifier within a high-risk background, driving various neurodevelopmental outcomes while allowing for the manifestation of disease even in the absence of the primary rare variant.

Most cases of *CHD2* related illness arise as *de novo* mutations often presenting early in life as epileptic encephalopathy. There have been reports of autosomal inheritance causing haploinsufficiency with a variety of clinical presentations from epilepsy in early childhood, to ASD in childhood, to schizophrenia, both childhood onset and more typical teen to adult onset. Poisson et al. presented a childhood-onset schizophrenia case associated with a mutation in *CHD2* at chr15:93,024,195 G-T (GRCh38) (17). Family A presents with a different extremely rare predicted LoF variant at chr15:93024653 C-A that is present in nearly all members sequenced from 3 generations. The sequenced carriers in generations 2 and 3 present with schizophrenia. A second generation family member is a parent of a third generation family member and is an obligate carrier though not sequenced and carries the diagnosis of schizophrenia. We hypothesize that especially high-density multiplex families, like Family A, are an enriched source of rare highly penetrant mutations and provide an important resource for studying the downstream biologic consequences. Further, given the shared features of the phenotypes presented we hypothesize that the biologic consequences of a variety of different such variants may converge.

Various mutations in *CHD2* are likely to have divergent molecular effects, depending on the position affected (18). *CHD2* is a member of the chromodomain helicase gene family, which has a variety of functions, including altering chromatin accessibility at sites where CHD proteins interact with transcription factors. Minor changes of missense or short truncations, such as the mutation in Family A that shortens the protein from 1,828 to 1,811 amino acids, are likely to have distinct effects based on changes in factor binding or effector domains. Furthermore, the effects of minor changes in CHD2 protein sequence would be expected to be modulated by interactions with other variant protein or mutated binding sequences to cause individual changes in penetrance. Minor alterations within a broadly interacting protein like CHD2 could underlie broad differences in diagnosis.

## Future Directions

These findings suggest a shared genetic architecture across various severe mental illnesses and demonstrate the utility of investigating rare, large effect variants that are identical by descent. Given the potential for founder effects among apparently unrelated PIC study families, some of our study families may share extremely rare mutations. We intend to sequence all remaining family members. Our future research will prioritize high-interest candidate variants to test whether these rare mutations dysregulate gene sets and may converge on shared downstream pathways. Further, given the nature of this population, the carriers of these identical rare mutations also share environmental risks. This presents a unique opportunity to study multiple family members at a cellular level. Our goal is to identify novel targets for therapeutic intervention that may have utility in the treatment of a broad range of patients suffering from serious mental illnesses.

## Methods

### Ascertainment and assessment

Families were identified through systematic screening of clinicians, treatment facilities, social services, and detailed family interviews. In the Azores, all four psychiatric hospitals and both general hospitals participated in the study. In Madeira, both psychiatric hospitals and the general hospital were involved. All participants provided informed consent, and the collection of blood samples and family history information was approved by the appropriate institutional review boards.

Diagnoses were based on data obtained using the Diagnostic Interview for Genetic Studies (DIGS) (19). Interviews were conducted at the participant’s clinical care site, in the home, or at another location of the participant’s choosing. Interviewers were highly trained psychiatrists, psychologists, or social workers.

“Interrater reliability was ensured through rigorous training and evaluation of interviewers prior to study initiation and annually thereafter. Reliability was periodically assessed through independent evaluations of 12 subjects by interviewers from different study sites, as well as by a senior supervisor. Chance-corrected interrater agreement (κ) was high for diagnoses of schizophrenia, bipolar disorder, and schizoaffective disorder (κ = 0.90–0.94). Comprehensive clinical narratives were completed for all subjects.” (9)

Best-estimate diagnoses, based on the Diagnostic and Statistical Manual of Mental Disorders, Fourth Edition and fifth editions (DSM-IV and V), were assigned by two independent blinded researchers after reviewing clinical information, the DIGS, the Operational Criteria Checklist for Psychotic Illness, and written narratives (20). Cases with discrepant diagnoses were reviewed by a third senior psychiatrist who was blinded to case status (21–23).

### Whole Genome Sequencing (WGS)

The individuals in this pilot study were sequenced along with participants in our inPSYght study (24). Individuals were whole genome sequenced (mean ± SD depth 26.8 ± 5.5).

### Preparation of libraries for cluster amplification and sequencing

(As per Broad protocol) An aliquot of genomic DNA (350ng in 50μL) was used as the input into DNA fragmentation (also known as shearing). Shearing was performed acoustically using a Covaris focused-ultrasonicator, targeting 385bp fragments. Following fragmentation, additional size selection was performed using a SPRI cleanup. Library preparation was performed using a commercially available kit provided by KAPA Biosystems (KAPA Hyper Prep without amplification module, product KK8505), and with palindromic forked adapters with unique 8-base index sequences embedded within the adapter (purchased from Roche). Following sample preparation, libraries were quantified using quantitative PCR (kit purchased from KAPA Biosystems), with probes specific to the ends of the adapters. This assay is automated using Agilent’s Bravo liquid handling platform. Based on qPCR quantification, libraries are normalized to 2.2nM and pooled into 24-plexes.

### Cluster amplification and sequencing (HiSeq X)

Sample pools were combined with HiSeqX Cluster Amp Regents EPX1, EPX2 and EPX3 into single wells on a strip tube using the Hamilton Starlet Liquid Handling system. Cluster amplification of the templates was performed according to the manufacturer’s protocol (Illumina) with the Illumina cBot. Flowcells were sequenced on HiSeqX utilizing sequencing-by-synthesis kits to produce 151bp paired-end reads. Output from Illumina software was processed by the Picard data-processing pipeline to yield CRAM or BAM files containing demultiplexed, aggregated aligned reads. All sample information tracking was performed by automated LIMS messaging.

### Quality control, functional annotation, and rare variant carrier identification

Whole-genome sequencing data for 359 samples (PIC samples were run with other samples) were imported from a joint-called VCF into a Hail MatrixTable, yielding an initial set of 31,445,428 variants. Genotype-level quality control was applied first: haploid calls on sex chromosomes were recoded as homozygous diploid to ensure uniform ploidy representation, and genotypes failing depth (DP) < 10, allele balance (AD) > 0.2, or genotype quality (GQ) > 20 thresholds were set to missing. Variant-level filtering focused on PASS-flagged variants, a call-rate filter of ≥0, and the final exclusion of sites overlapping low-complexity regions (LCRs) and segmental duplications (SegDup), retaining 19,258,933 high-quality variants (61.2% of the original input). Sex was imputed for every sample using depth-normalized X/Y ploidy ratios and the non-PAR X-chromosome inbreeding coefficient (F-statistic), yielding 200 female, 155 male, and 4 ambiguous calls, with 94.4% concordance against recorded metadata. Population structure was characterized by projecting study samples onto a reference PCA space constructed from 3,223 HGDP 1K Genomes samples. A Random Forest classifier trained on the top five PCs assigned 99.7% (min_prob = 0.70) of samples to European ancestry, consistent with the predominantly Iberian-like genetic background of this cohort.

The QC-filtered dataset was restricted to coding sequence variants plus 50 bp of flanking intronic sequence (CDS+50 bp), leaving 283,504 variants. All variants were annotated with allele frequencies from external references, gnomAD and UK Biobank. Functional consequences were annotated for all variants using the Variant Effect Predictor (VEP) on canonical transcripts, classifying each as stop-gained, frameshift, essential splice-site, missense, synonymous, or other. We focused on variants with AF < 1%, identifying 139,006 rare coding variants.

Rare coding variants were intersected with a panel of curated neuropsychiatric and neurodevelopmental gene sets: schizophrenia (SCHEMA), autism and broader developmental disorders (ASD/DD/NDD), and gnomAD pLI-constrained. Loss-of-function (LoF) variants were classified in two ways: LoFHC retained only high-confidence predicted LoF variants as defined by LOFTEE (stop-gained, essential splice-site, frameshift passing LOFTEE filters), while LoFall included all predicted LoF annotations regardless of LOFTEE confidence. This gene set served as the primary resource for studying the segregation of familial rare variants.

## Data Availability

All data produced in the present study are available upon reasonable request to the authors

## Author Contributions

**Conceptualization:** Carlos and Michele Pato; **Methodology:** Carlos N Pato, Michele T. Pato, Jennifer Mulle, Ronald P. Hart, Zhiping Pang, James A. Knowles, Tarjinder Singh, Priya Maddhesiya, Celia Carvalho, Alison Merikangas, Helena Medeiros, Tim B. Bigdeli, Hamed Kazemi, John Drake, Vladimir I. Vladimirov, Brion S. Maher, Silviu-Alin Bacanu, Ben Neale, Ayman Fanous; **Formal Analysis:** Carlos N Pato, Michele T. Pato, Jennifer Mulle, Ronald P. Hart, Zhiping Pang, James A. Knowles, Tarjinder Singh, Priya Maddhesiya, Celia Carvalho, Alison Merikangas, Helena Medeiros, Tim B. Bigdeli, Hamed Kazemi, John Drake, Vladimir I. Vladimirov, Brion S. Maher, Silviu-Alin Bacanu, Ben Neale, Ayman Fanous **Investigation:** Carlos N Pato, Michele T. Pato, Jennifer Mulle, Ronald P. Hart, Zhiping Pang, James A. Knowles, Tarjinder Singh, Priya Maddhesiya, Celia Carvalho, Alison Merikangas, Helena Medeiros, Tim B. Bigdeli, Hamed Kazemi, John Drake, Vladimir I. Vladimirov, Brion S. Maher, Silviu-Alin Bacanu, Ben Neale, Ayman Fanous^;^ **Resources (PIC Cohort):** Carlos and Michele Pato; **Data Curation:** Carlos N Pato, Michele T. Pato, Jennifer Mulle, Ronald P. Hart, Zhiping Pang, James A. Knowles, Tarjinder Singh, Priya Maddhesiya, Celia Carvalho, Alison Merikangas, Helena Medeiros, Tim B. Bigdeli, Hamed Kazemi, John Drake, Vladimir I. Vladimirov, Brion S. Maher, Silviu-Alin Bacanu, Ben Neale, Ayman Fanous **Writing – Original Draft:** Carlos and Michele Pato; **Writing – Review & Editing:** Carlos N Pato, Michele T. Pato, Jennifer Mulle, Ronald P. Hart, Zhiping Pang, James A. Knowles, Tarjinder Singh, Priya Maddhesiya, Celia Carvalho, Alison Merikangas, Helena Medeiros, Tim B. Bigdeli, Hamed Kazemi, John Drake, Vladimir I. Vladimirov, Brion S. Maher, Silviu-Alin Bacanu, Ben Neale, Ayman Fanous; **Visualization:**Carlos and Michele Pato; **Supervision:** Carlos and Michele Pato, Ayman Fanous, Ben Neale; **Project Administration:** Carlos and Michele Pato, Ayman Fanous, Ben Neale; **Funding Acquisition:** Carlos and Michele Pato, Ayman Fanous, Ben Neale.

## Acknowledgements

1. M. Helena Azevedo, Carlos Paz Ferreira, Antonio Macedo, Jose Valente.

